# Contribution of Marijuana Legalization to the U.S. Opioid Mortality Epidemic: Individual and Combined Experience of 27 States and District of Columbia

**DOI:** 10.1101/19007393

**Authors:** Archie Bleyer, Brian Barnes

## Abstract

**Background:** Prior studies of U.S. states as of 2013 and one state as of 2015 suggested that marijuana availability reduces opioid mortality (marijuana protection hypothesis). This investigation tested the hypothesis with opioid mortality trends updated to 2017 and by evaluating all states and the District of Columbia (D.C.).

**Methods:** Opioid mortality data obtained from the U.S. Centers for Disease Control and Prevention were used to compare opioid death rate trends in each marijuana-legalizing state and D.C. before and after medicinal and recreational legalization implementation and their individual and cumulative aggregate trends with concomitant trends in non-legalizing states. The Joinpoint Regression Program identified statistically-significant mortality trends and when they occurred.

**Results:** Of 23 individually evaluable legalizing jurisdictions, 78% had evidence for a statistically-significant acceleration of opioid death rates after medicinal or recreational legalization implementation at greater rates than their pre-legalization rate or the concurrent composite rate in non-legalizing states. All four jurisdictions evaluable for recreational legalization had evidence (p <0.05) for subsequent opioid death rate increases, one had a distinct acceleration, and one a reversal of prior decline. Since 2009-2012, when the cumulative-aggregate opioid death rate in the legalizing jurisdictions was the same as in the non-legalizing group, the legalizing group’s rate accelerated increasingly faster (p=0.009). By 2017 it was 67% greater than in the non-legalizing group (p <<0.05).

**Conclusions:** The marijuana protection hypothesis is not supported by recent U.S. data on opioid mortality trends. Instead, legalizing marijuana appears to have contributed to the nation’s opioid mortality epidemic.

## Introduction

According to the Institute for Health Metrics and Evaluation, the United States (U.S.), leads the world in opioid death rate by more than twice that of next closest county [Libya]) and is 2^nd^ worldwide in cannabis use disorder prevalence (Fig. 1).^1^ Are these two dire statistics related, and if so, how?

**Figure 1.**
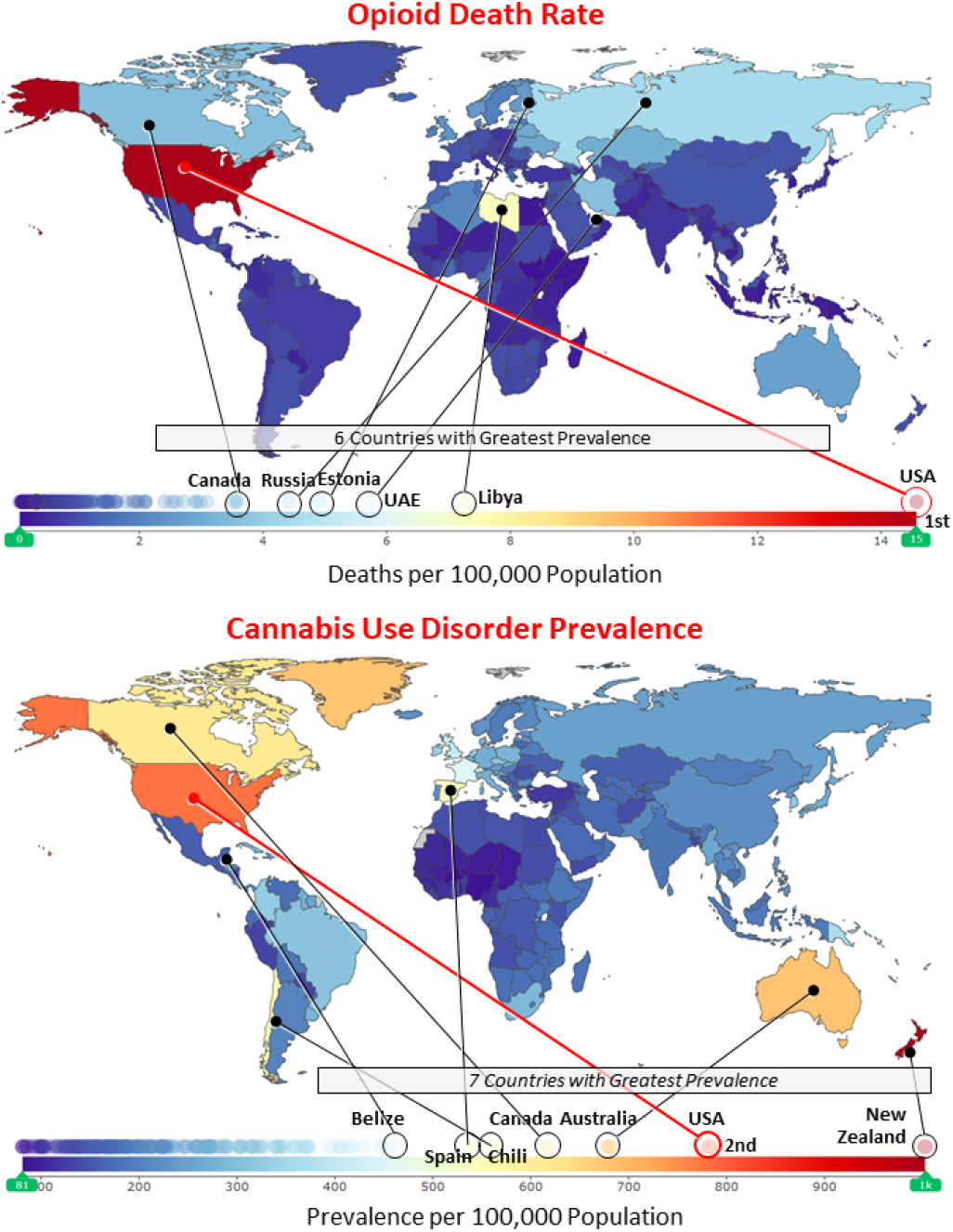
**World Ranking** (of 195 Countries and Territories) **of Opioid Death Rate** (upper panel) **and Cannabis Use Disorder** (lower panel), **2017**. <u>Data Source</u>: Institute for Heath Metrics and Evaluation^2^

According to data at the U.S. Centers for Disease Control and Prevention,^2^ the U.S.’s national opioid death rate trend since 1999 is directly proportional to the percent of American’s able to access marijuana legally (p <10^−10^, Fig. 2). To what extent is this correlation cause and effect (marijuana use leading to opioid abuse)?

**Figure 2:**
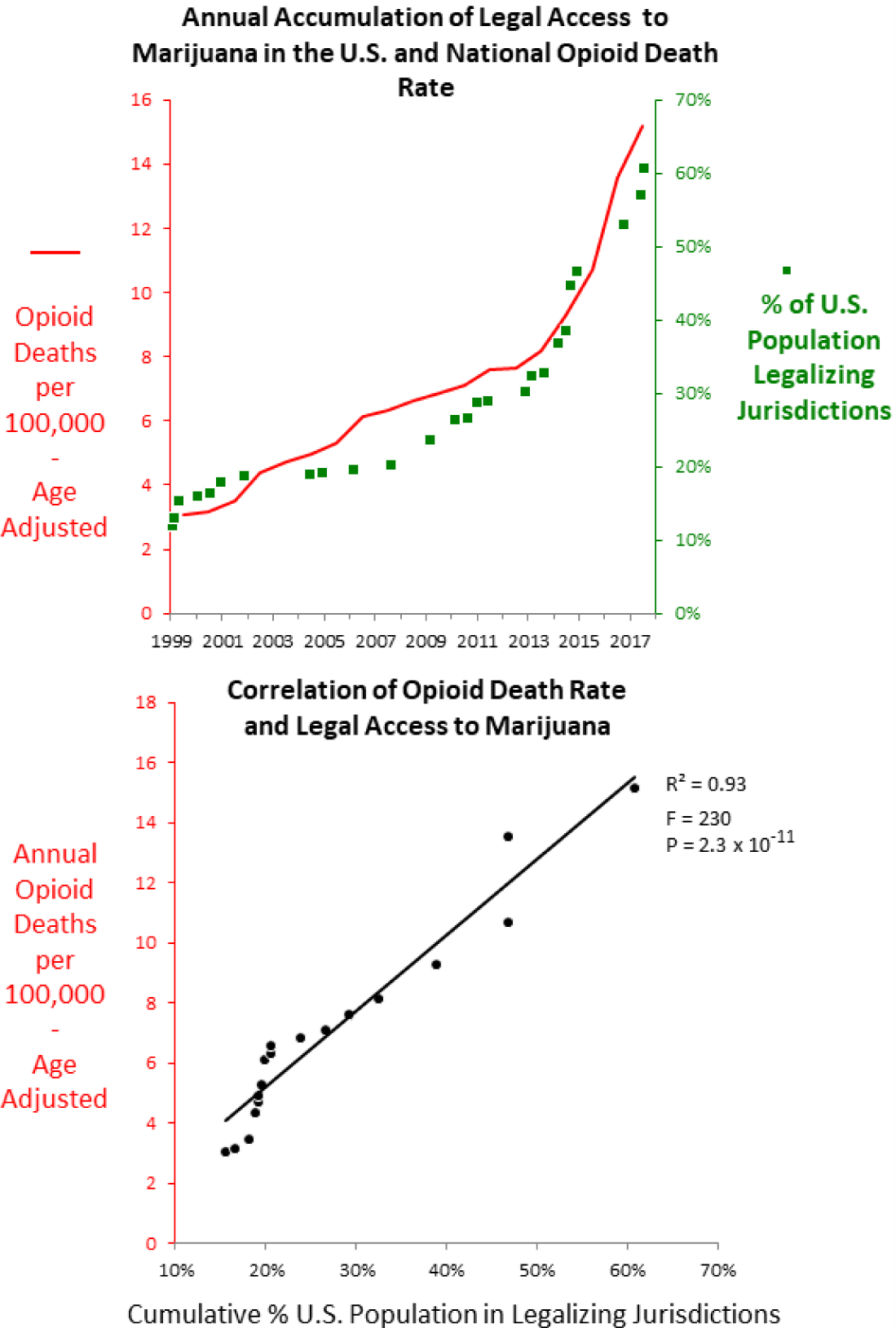
Annual Cumulative % of U.S. Population in Marijuana Legalizing Jurisdictions and Opioid Death Rates in the U.S., 1999-2017. <u>Data Source</u>: CDC WONDER^2^

Three prior reports^3-5^ from the U.S. presented data supporting the marijuana protection hypothesis: *availability of marijuana reduces deaths from opioids*. Bachhuber et al^3^ analyzed opioid death rates as of 2010 after medicinal marijuana legalization in 10 U.S. states. Powell et al^4^ extended the opioid-related mortality analysis to 2013 and added 6 states and the District of Columbia (D.C.) that legalized medicinal marijuana during 2010-2013. Livingston et al^5^ investigated opioid deaths after recreational legalization in a single state, Colorado, as of December 2015, 24 months after legalization. These authors concluded that cannabis legalization was associated with significantly lower state-level opioid mortality rates^3-5^ and that the reduction represented a reversal of opioid-related death trends.^5^ Although neither of the first two reports were claimed to be definitive, editorial commentary^6,7^ and the public response^8^ generally accepted the results as supportive of legalization. Billboards in major U.S. metropolitan communities subsequently claimed “states that legalized marijuana had 25% fewer opioid related deaths”, with reference to the Bachhuber report.^9^

We undertook our study to more adequately test the marijuana protection hypothesis by evaluating all 50 states and D.C., adding six more states that subsequently legalized marijuana, extending the follow-up of all legalizing jurisdictions by four more years, and including the effect of recreational marijuana legalization in three more states. We tested the hypothesis via three comparisons: each legalizing jurisdiction before and after legalization implementation; each legalizing jurisdiction relative to concomitant trends in all non-legalizing states; and the cumulative aggregate of legalizing jurisdictions with concomitant trends in non-legalizing jurisdictions. The results strengthen a preliminary and partially inaccurate correspondence^10^ and do not support the marijuana protection hypothesis.

## Methods

Mortality data were obtained from the Centers for Disease Control and Prevention (CDC), National Center for Health Statistics CDC WONDER Multiple Cause of Death Files^2^ that provide death data from 1999 to, as of this analysis, 2017. All annual death rates obtained from CDC WONDER were age-adjusted according to the United States 2000 standard population. Rarely, when the CDC database provided the number of deaths and population but not the corresponding death rate, the rate was calculated from the deaths and population data.

Trend analysis was performed with the Joinpoint Regression Program, version 4.6.0.0.11 The Joinpoint Regression Program identifies trend inflections (“joinpoints”) to determine when a trend changes to another trend, the probability range of the inflection, and the average annual percent change (AAPC) and p values for each trend detected. It allows statistical significance testing of trend differences and does not depend on parallel trend assumptions. Joinpoint analysis was applied with weighted least squares, Poisson methods, logarithmic transformation, and standard errors.

International Classification of Disease (ICD) T Codes used for the primary analysis were T40.0 opium, T40.1 heroin, T40.2 other opioids, T40.3 methadone, T40.4 other synthetic opioids, and T40.6 other synthetic narcotics. The opiates include morphine, hydromorphone, oxycodone, fentanyl, semisynthetic fentanyl moieties, heroin, opium, codeine, meperidine, methadone, propoxyphene, tramadol, and other/unspecified narcotics. The T categories were applied in conjunction with the following ICD codes: X40-X44 accidental poisoning, X60-X64 intentional self-poisoning, Y10-Y14 other poisoning.

Table 1 lists each state and D.C. by whether and when marijuana legalization for medicinal or recreational use was implemented. The implementation dates for jurisdictions that legalized before 2015 are those published by Powell et al^4^ and those since were obtained from other sources.^12,13^ The earliest available opioid mortality data on the CDC website is January 1999. One state (California) implemented marijuana legalization 26 months before the first available opioid mortality data (Table 1). The two most recent jurisdictions to legalize marijuana during before December 2017, Ohio and Pennsylvania, did so in April and June of 2016, respectively, but did not implement via state-approved dispensaries for more than a year.

**Table 1.** State and D.C. Marijuana Use Legalization Status and Legalization Implementation Date.

| State and D.C. | Use Legalization | Legalization Implementation Date† |  | Evaluability for Trend Correlation with Available Opioid Death Data |
| --- | --- | --- | --- | --- |
|  |  | Medicinal | Recreational |  |
| Legalizing States and D.C. |  |  |  |  |
| 1 California | Medicinal and recreational | 11/5/1996³ | 1/1/2018 | Medicinal only but limited* |
| 2 Oregon | Medical and recreational | 12/3/1998⁴ | 1/1/2017 | Medicinal only |
| 3 Washington | Medicinal and recreational | 12/3/1998³ | 12/6/2012 | Medicinal & recreational |
| 4 Alaska | Medicinal and recreational | 3/4/1999³ | 2/24/2015 | " |
| 5 Maine | Medicinal and recreational | 12/23/1999³ | 5/1/2018 | Medicinal only |
| 6 Hawaii | Medicinal only | 6/16/2000³ |  | " |
| 7 Colorado | Medicinal and recreational | 12/28/2000³ | 11/6/2012‡ | Medicinal & recreational |
| 8 Nevada | Medicinal and recreational | 10/1/2001³ | 7/1/2017 | Medicinal only |
| 9 Vermont | Medicinal only | 7/1/2004³ |  | " |
| 10 Montana | Medicinal only | 11/2/2004³ |  | " |
| 11 Rhode Island | Medicinal only | 1/3/2006³ |  | " |
| 12 New Mexico | Medicinal only | 7/1/2007³ |  | " |
| 13 Michigan | Medicinal only | 12/4/2008³ |  | " |
| 14 New Jersey | Medicinal only | 6/1/2010³ |  | " |
| 15 D.C. | Medicinal and recreational | 7/27/2010³ | 2/26/2015 | Medicinal & recreational |
| 16 Arizona | Medicinal only | 11/29/2010³ |  | " |
| 17 Delaware | Medicinal only | 5/13/2011³ | 2017 | Medicinal only |
| 18 Connecticut | Medicinal only | 10/1/2012³ |  | " |
| 19 Massachusetts | Medicinal and recreational | 1/1/2013³ | 7/2018 | " |
| 20 New Hampshire | Medicinal only | 7/23/2013³ |  | " |
| 21 Illinois | Medicinal only | 1/1/2014³ |  | " |
| 22 Minnesota | Medicinal only | 5/30/2014³ |  | " |
| 23 New York | Medicinal only | 7/1/2014³ |  | " |
| 24 Maryland | Medicinal only | 6/1/2014** |  | " |
| 25 Pennsylvania | Medicinal only | 4/17/2016 |  | " |
| 26 Ohio | Medicinal only | 6/8/2016 |  | " |
| Non-Legalizing States |  |  |  |  |
| 1 Georgia | Medicinal only | 4/16/2015 |  | Minimal legalization^ Evaluable |
| 2 Florida | Medicinal only | 11/2016 |  | End of 2016; Fully Evaluable |
| 3 Arkansas | Medicinal only | 11/8/2016 |  | " |
| 4 North Dakota | Medicinal only | 11/8/2016 |  | " |
| 5 West Virginia | Medicinal only | 7/2018 |  | Fully Evaluable |
| 6 Louisiana | Medicinal only | 2018^^ |  | " |
| 7 Alabama |  |  |  | " |
| 8 Idaho |  |  |  | " |
| 9 Indiana |  |  |  | " |
| 10 Iowa |  |  |  | " |
| 11 Kansas |  |  |  | " |
| 12 Kentucky |  |  |  | " |
| 13 Mississippi |  |  |  | " |
| 14 Missouri |  |  |  | " |
| 15 Nebraska |  |  |  | " |
| 16 North Carolina |  |  |  | " |
| 17 Oklahoma |  |  |  | " |
| 18 South Carolina |  |  |  | " |
| 19 South Dakota |  |  |  | " |
| 20 Tennessee |  |  |  | " |
| 21 Texas |  |  |  | " |
| 22 Utah |  |  |  | " |
| 23 Virginia |  |  |  | " |
| 24 Wisconsin |  |  |  | " |
| 25 Wyoming |  |  |  | " |

<sup>†</sup>Source either Powell et al.<sup>3</sup> or Wikipedia<sup>12,13</sup> \*Legalization implementation >2 years before 1<sup>st</sup> available opioid mortality data
<sup>‡</sup>2009 rulings enabled widespread use<sup>15</sup> \*\*2003 legislation enabled in 2014<sup>3</sup> ^CBD oil for epilepsy ^^2015 legislation enabled in 2018

Each legalizing jurisdiction was evaluated individually for opioid mortality trends before and after legalization implementation, if feasible, and in combination with other legalizing jurisdictions in a comparison of all legalizing and non-legalizing jurisdictions (composite analysis).

### Legalizing Jurisdictions

Four states were inevaluable for individual trend analysis since they either implemented legalization too early (California) before the available opioid mortality data or too recently (Florida, Ohio, Pennsylvania) to assess their post-implementation opioid mortality trend (Table 1). Excluding these four states, a total of 22 states and D.C. were each evaluable for trend analysis (Table 1).

For individual jurisdictions (state and D.C.) that legalized marijuana, two comparisons were conducted: 1) the opioid death rate trends before and after legalization implementation and 2) the post-legalization opioid death trend and its statistically significant AAPCs derived with Joinpoint Regression Program^11^ was compared with the composite of non-legalizing states.

### Non-Legalizing Jurisdictions

The non-legalizing group consisted of 24 states that had not implemented legalization by December 2017. Georgia was included in the non-legalizing group since it legalized only CBD oil and only for epilepsy.

The composite of legalizing and non-legalizing groups of jurisdictions were compared with the former aggregated by accumulation of each legalizing jurisdiction when it implemented legalization during 1999 2017, after either medicinal or recreational legalization implementation. Given the relatively few states that legalized marijuana during the early years—nine states during 1996-2007—the comparison between the legalizing and non-legalizing groups was regarded not to be meaningful until either the legalizing group rate was stable or both groups has the same rates, both of which occurred during 2009-2011. Because the rates were identical in 2009-2011 and had overlapping 95% confidence intervals in 2012, difference-in-difference methodology was unnecessary to compare the subsequent trends. Because of delays in implementing medicinal legalization, the two most recent legalizing states, Pennsylvania and Ohio, were both evaluated with, and excluded from, the composite legalizing group.

## Results

Analyses of individual legalization jurisdictions compared with a composite result of non-legalizing jurisdictions are presented first, followed by comparisons of a composite of legalizing jurisdictions with a composite of non-legalizing jurisdictions.

### Individual Jurisdiction (State and D.C.) Analysis

Figure 3 provides the guide to Figures 4, 5 and 6 that depict the annual opioid death rate during 1999-2017 in each of the individually evaluable 22 legalizing states and D.C. The green and black curves are the 95% confidence intervals for the legalizing and non-legalizing jurisdictions, respectively. The red and green vertical bars indicate medicinal and recreational legalization implementation, respectively. Each legalizing jurisdiction was assessed both for its opioid death trend compared to the non-legalizing group and for its trend before and after legalization implementation.

**Figure 3.**
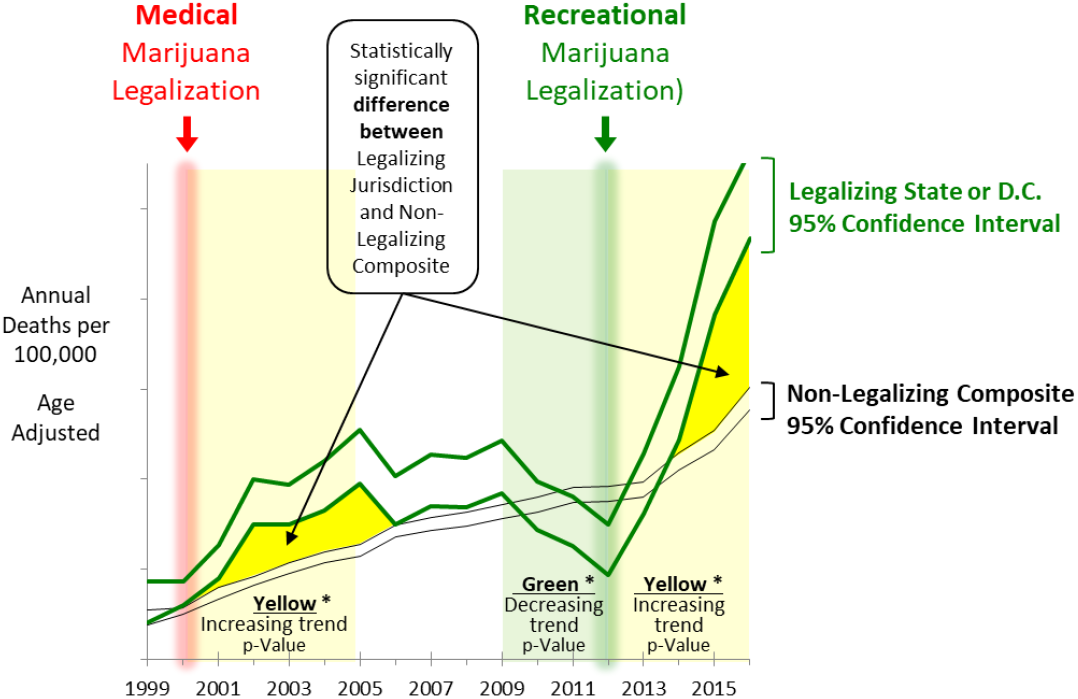
Guide to Figures 4-7. **Green curves:** 95% confidence interval for the state or D.C. that legalized marijuana **Black curves:** 95% confidence interval for the 25 States that as of December 2016 had not implemented marijuana legalization. **Vertical red and green bars:** implementation of medicinal and recreational legalization, respectively. \***Light Yellow and green backgrounds:** trend change for the legalizing state (or D.C.) identified by, and p-value provided by, joinpoint analysis11 of annual rates. **Dark yellow areas**: statistically-significant increases compared to non-legalizing group.

**Figure 4.**
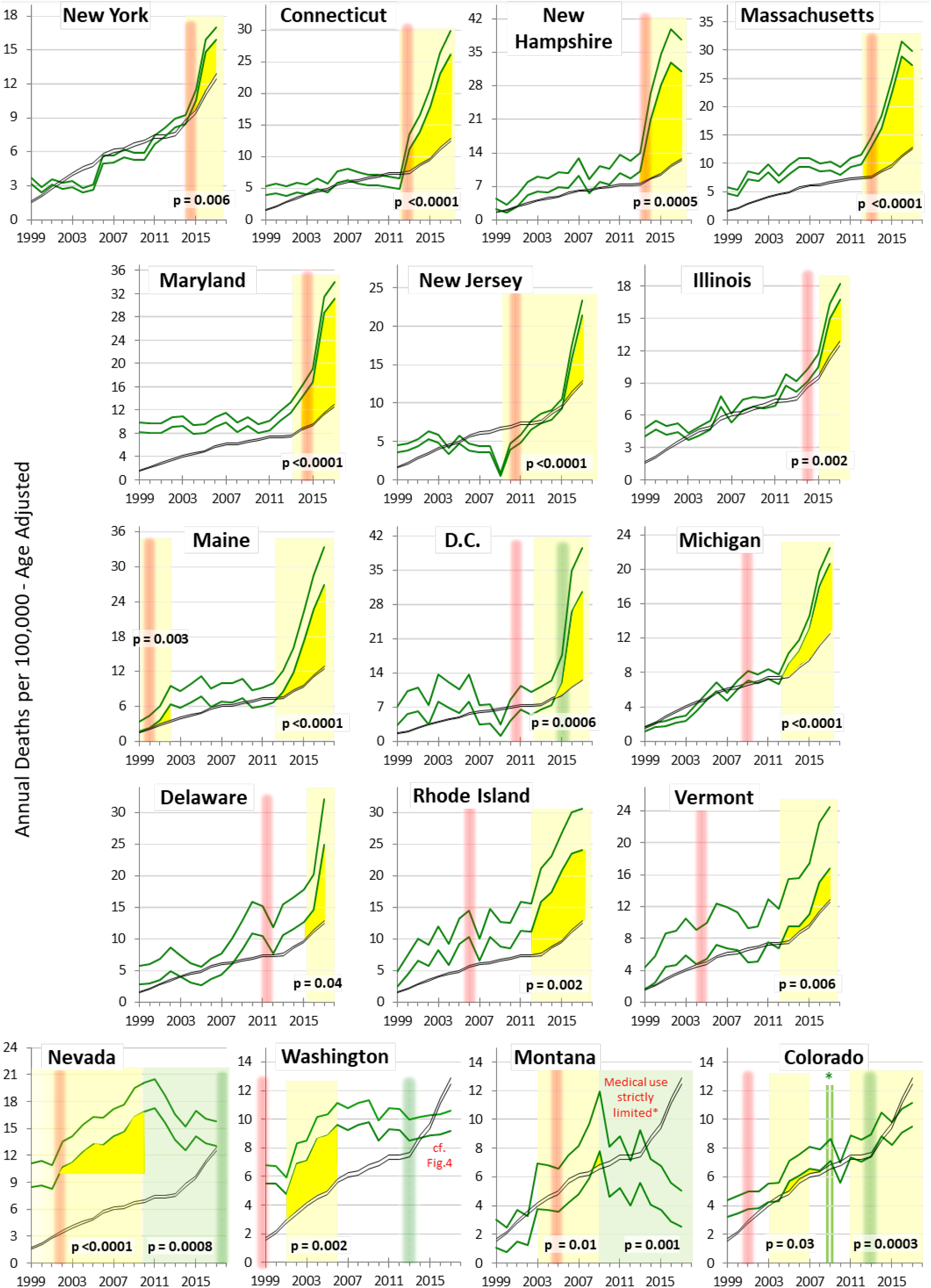
95% Confidence Intervals of Annual Opioid Death Rate in Legalizing Jurisdictions with Acceleration of Their Death Rate after Legalization Implementation (green curves) and in Non-Legalizing States Composite (black curves). The jurisdictions are arrayed in order of temporal association of legalization implementation with statistically-significant increase of their subsequent death rate, except for the bottom row which is more multifaceted. The guide to the curves, vertical red and green lines, p-values, background highlighting, and yellow areas is in Figure 3. *In Colorado, several rulings allowed widespread use of marijuana with store front dispensaries across the state and the “Colorado Green Rush”.^15^ <u>Data Source</u>: CDC WONDER^2^,

**Figure 5.**
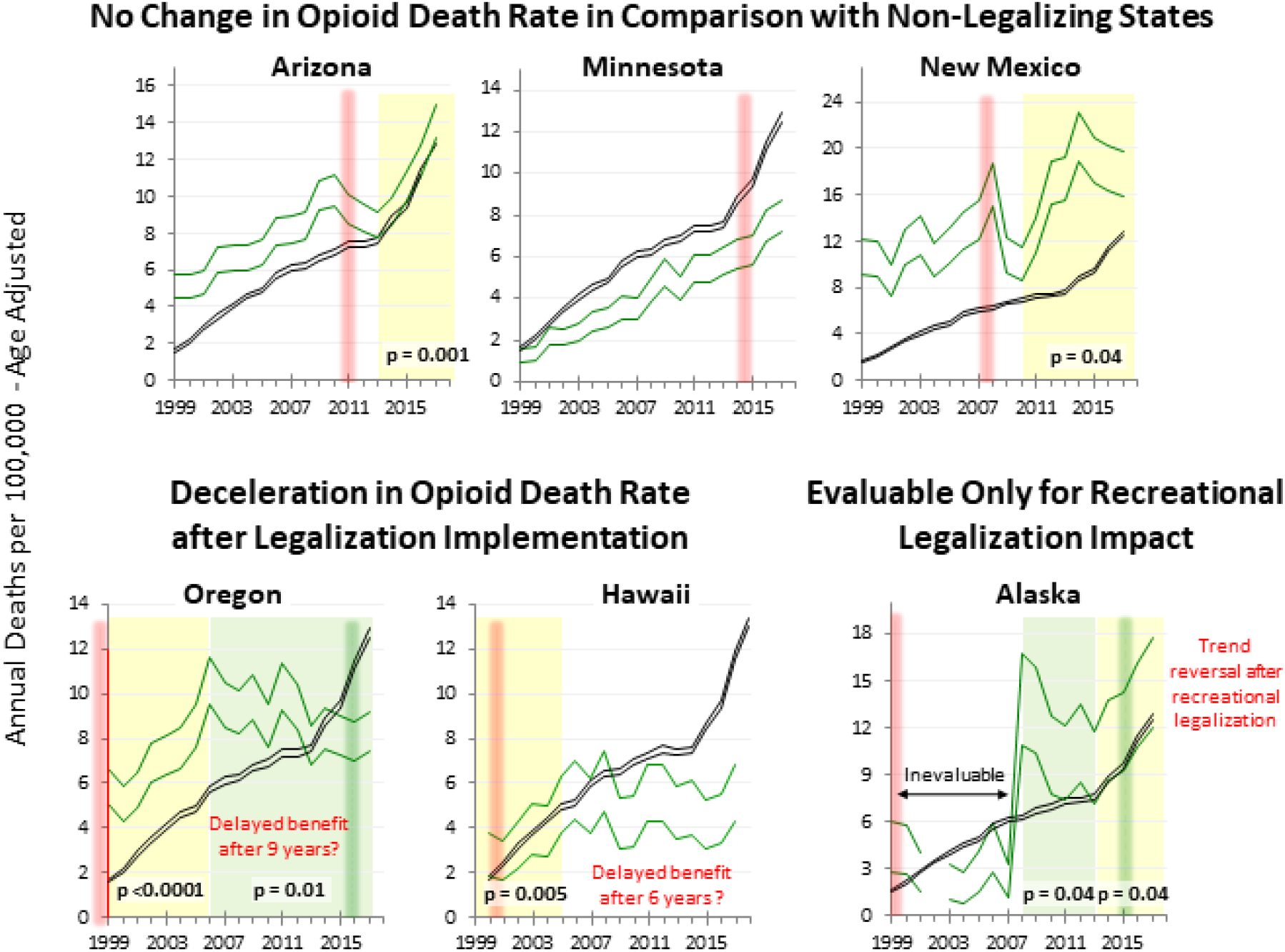
States with Either No Apparent Change, Delayed Deceleration, or Evaluable Only for Recreational Legalization. (green data). The guide to the curves, vertical lines, joinpoint analysis input, p values, and background highlighting is in Figure 3. Data Source: CDC WONDER ^2^

**Figure 6:**
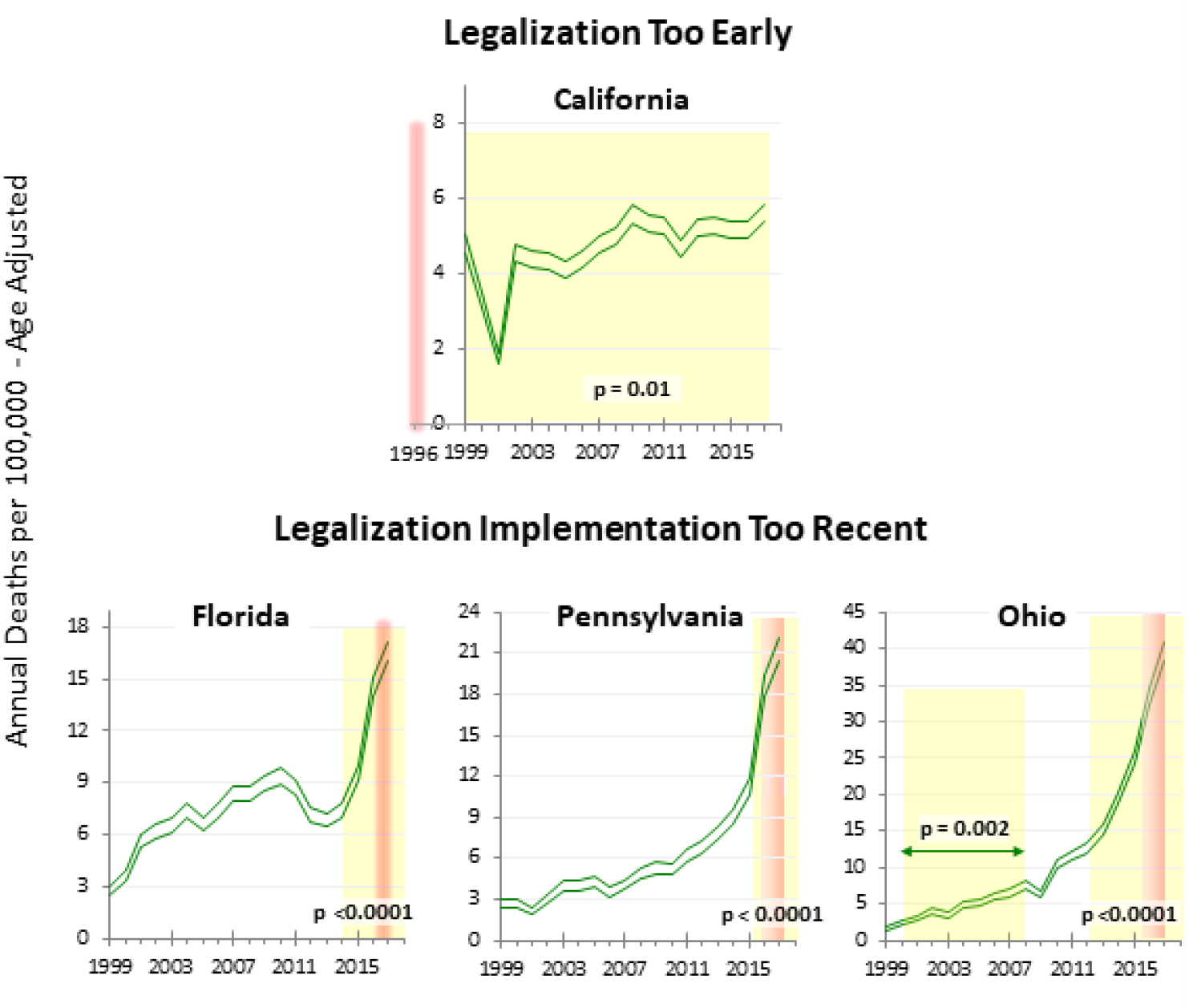
95% Confidence Intervals of Annual Opioid Death Rate in Four States Not Evaluable for Impact on Opioid Mortality after Marijuana Legalization Implementation. The guide to the curves, red bars, joinpoint analysis input, p-values, and background highlighting is in Figure 3. <u>Data Source:</u> CDC WONDER^2^

### Comparison with Non-Legalizing States

Of 23 jurisdictions individually evaluable for comparison with the non-legalizing group of jurisdictions (Figs. 4 and 5), 17 (74%) had statistically-significantly higher opioid death rates after legalization implementation than the non-legalizing group (Fig. 4), as indicated by separation of the 95% C.I. zones between the legalizing jurisdiction and the non-legalizing group (p <0.05). Fifteen (88%) of the 17 had multiple 95% C.I. separations between them and the non-legalizing group (p <<0.05). Montana had the least degree of separation but the trend thereafter abruptly reversed after medicinal marijuana access in the state was severely limited and the number of medical cardholders plummeted from 31,000 in May 2014 to 9,000 six months later.^14^ Three states (Arizona, Minnesota, New Mexico) had trends that were similar to the non-legalizing group (Fig. 5). Two states that had a delayed reduction in their annual opioid death rate ultimately had lower annual opioid death rates than the non-legalized group of states (Oregon 15 years later, Hawaii 7 years later) (Fig. 5). Alaska had acceleration of its opioid death rate after recreational legalization. that was preceded by 5 years of a declining rate (Fig. 5). In total, 18 (78%) of the evaluable legalizing jurisdictions have evidence for worsening of their opioid death rates than the non-legalizing group.

### Comparison of Pre- and Post-Legalization Trends

Four jurisdictions could not be individually compared for post- versus pre-legalization trend since they legalized too early (California) or too recently (Florida, Pennsylvania and Ohio) during the available 1999-2017 span of available data (Fig. 6). Of the evaluable 19 jurisdictions, 16 had acceleration after medicinal legalization, within 1 to 2 years in Connecticut, Illinois, Maine, Massachusetts, Maryland, Montana, Nevada, New Jersey, New Hampshire, and New York, and within 3 to 7 years in D.C., Delaware, Michigan, Rhode Island, Vermont and Washington. Maine’s acceleration began 2 years after medicinal legalization and again 10 years later. The three states (Arizona, Minnesota, New Mexico) cited above with trends similar to the non-legalizing group also did not appear to have a change in trend before and after legalization implementation. Two states (New Mexico, Arizona) had greater variability after legalization but ultimately assumed the pre-legalization trend. Two states (Oregon, Hawaii) appear to have had a trend reversal after 6 to 9 years

Figure 7 shows the opioid death rate trends for the four jurisdictions that legalized recreational use and were evaluable for comparison with their recreational pre-legalization trend. After recreational legalization, D.C had a striking acceleration of its opioid death rate (from AAPC of 2.16 (p=0.14) to 50.6 (p=0.004) and Alaska had reversal of what was a declining trend before recreational legalization to an AAPC of 12.0 (p=0.02). Colorado had an increase (AAPC 5.5, p <0.0003) the year after several statewide rulings resulted in widespread use of marijuana (“Colorado Green Rush”) with storefront dispensaries throughout the state and protection from federal intervention.15 Washington had no significant change, although it has had 4 consecutive years of increase (p=0.004) after a downward trend prior to recreational legalization.

**Figure 7.**
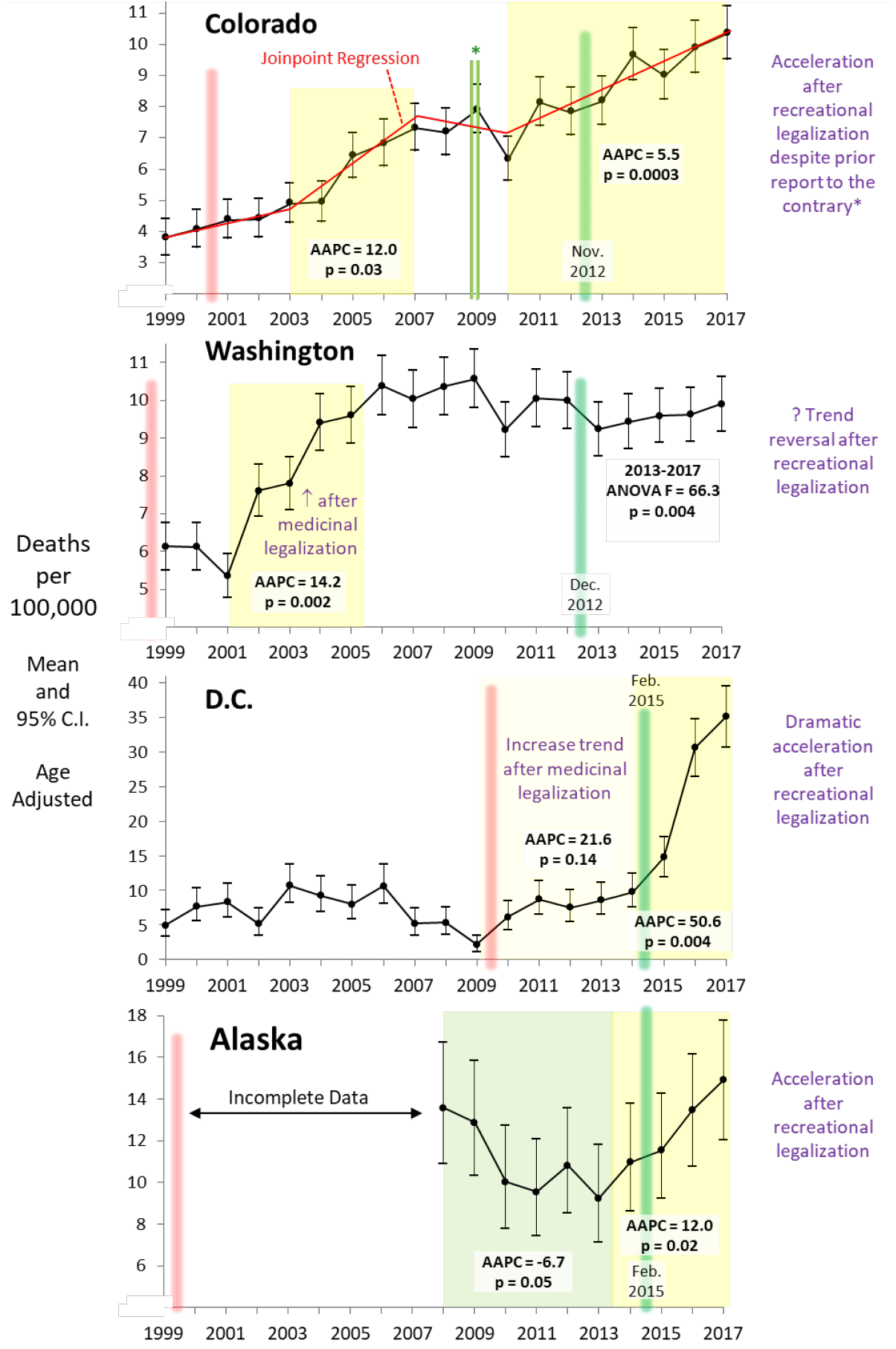
Mean and 95% C.I. of Annual Opioid Death Rates in Jurisdictions Evaluable for Impact of Recreational Marijuana Legalization. As per Figure 3, vertical bars designate legalization implementation for medicinal (red) and recreational (green) use and colored backgrounds indicate statistically-significant trends as determined by Joinpoint regression. *Colorado “Green Rush”^15^ is described in Figure 4 legend. <u>Data Source</u>: CDC WONDER^2^, Livingston et al.^5^

### Pre-Legalization and Non-Legalizing Comparisons

In summary, of the 23 evaluable jurisdictions, 17 states and D.C. (78%) had a statistically-significant acceleration of their opioid death rates after medicinal or recreational legalization at greater rates than the composite rate in non-legalizing states and/or their pre-legalization rate (Fig. 8). Three states had no change after legalization implementation in comparison with their pre-legalization trend or with the composite trend in non-legalizing states. Two states had deceleration of their opioid death rate after legalization implementation and in comparison with the non-legalizing states, but neither of these had, as of December 2017, statistically-significant lower rates after legalization implementation than before. Of four jurisdictions evaluable for recreational legalization, each had significant (p <0.05) evidence for subsequent increases in their opioid death rate and two had either a distinct acceleration or reversal of prior decline.

**Figure 8.**
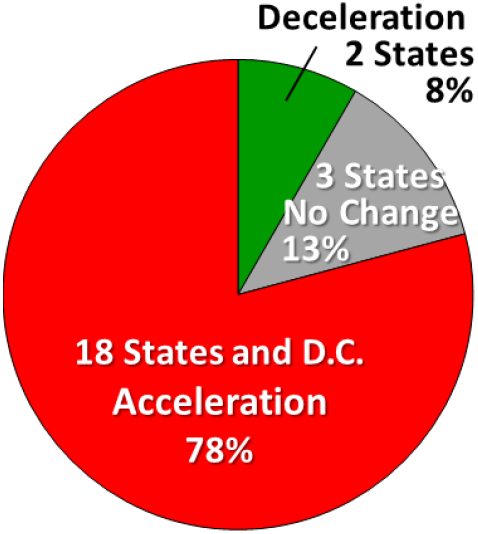
Summary of 23 Jurisdictions Evaluable for Opioid Death Rate Trend after Marijuana Legalization Implementation, by Trend Trajectory.

## Composite Analysis

Figure 9 shows the annual opioid death rates in legalizing and non-legalizing jurisdiction during 2009-2017, after 12 states had implemented (medicinal) legalization. During 2009-2012, the annual opioid death rate was the same in the non-legalizing jurisdictions and the cumulative aggregate of the legalizing jurisdictions. The rate increases during 2009-2012 also were the same (AAPC [p-value] = 2.1[0.33] and 3.6 [0.05], respectively). Thereafter, the death rate increased one year earlier and more rapidly in the legalizing group. The annual rate during 2015-2017 (AAPC = 24.5, p=0.04) was nearly twice that in the non-legalizing group (AAPC = 13.3, p=0.0001) and by 2017 it was 67% higher than in the non-legalizing group (p <<0.05). The difference between the legalizing and non-legalizing groups for all 9 years during 2009 2017 was statistically significant (p=0.009).

**Figure 9.**
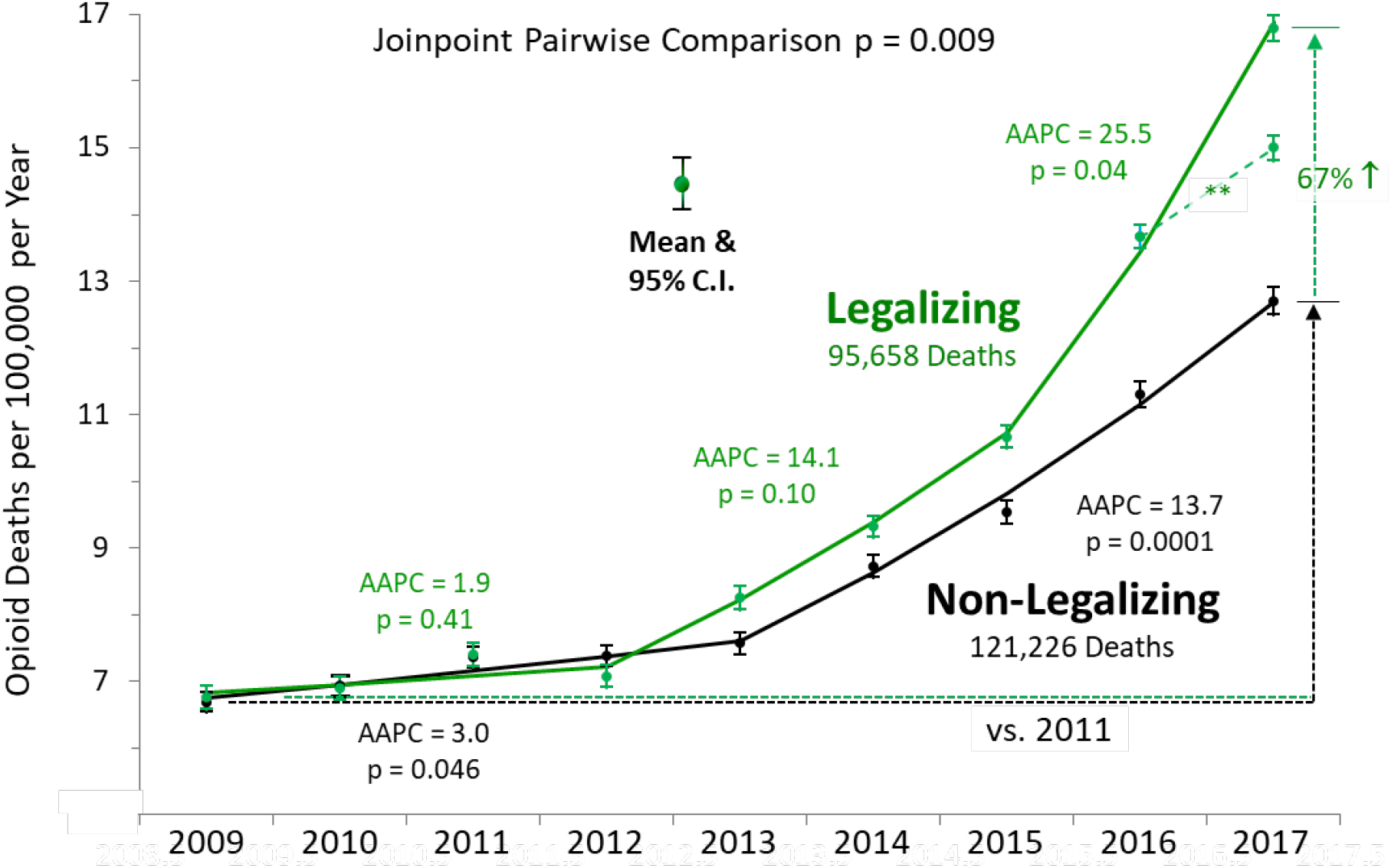
Joinpoint/AAPC* Analysis of Annual Opioid Death Rates in Non-Legalizing (24 states, black data) and Cumulative Aggregate of Legalizing Jurisdictions (26 states + D.C., green data) during 2009 2017. *AAPC – average annual percent change. **Excluding Pennsylvania and Ohio.<u>Data Source</u>: CDC WONDER^2^

From area-under-the-annual-curve analysis of joinpoint regressions, 71% of the total opioid death rate increase in the U.S. during 2009-2017 occurred in the legalizing jurisdictions. If Pennsylvania and Ohio were excluded from the legalizing group, given that they were the most recent states to legalize and had not fully implemented marijuana availability by the end of 2017, the increase from 2016 to 2017 in the legalizing group was less but still distinctly higher (37% greater increase as of 2017, p <<0.05, 63% of 2009-2017 total opioid death rate increase in legalizing jurisdictions).

## Discussion

<u>Note</u>: After this report was submitted for publication, investigators at Stanford University, The Network for Public Health Law, Carrboro, NC New York University, and Center for Innovation to Implementation, Veterans Affairs Health Care System, Palo Alto, has a report published in PNAS with supplementary data the yielded similar results:

The divergence in the death rates during 2012-2017 is similar to the separation we observed in Figure 9.

The primary result of their investigation was that association between medical cannabis laws and opioid overdose mortality reversed over time, from a reduction in opioid mortality to an increase that exceeded the opioid death rate prior to marijuana legalization. By 2017 the states that had legalized marijuana had a statistically significant greater increase in opioid death rate than they did before legalization:

The red outline in Figure 11 identifies a statistically significant greater opioid death rate than all previous evaluated years. Our analyses of their results support the findings of our investigation.

**Figure 10.**
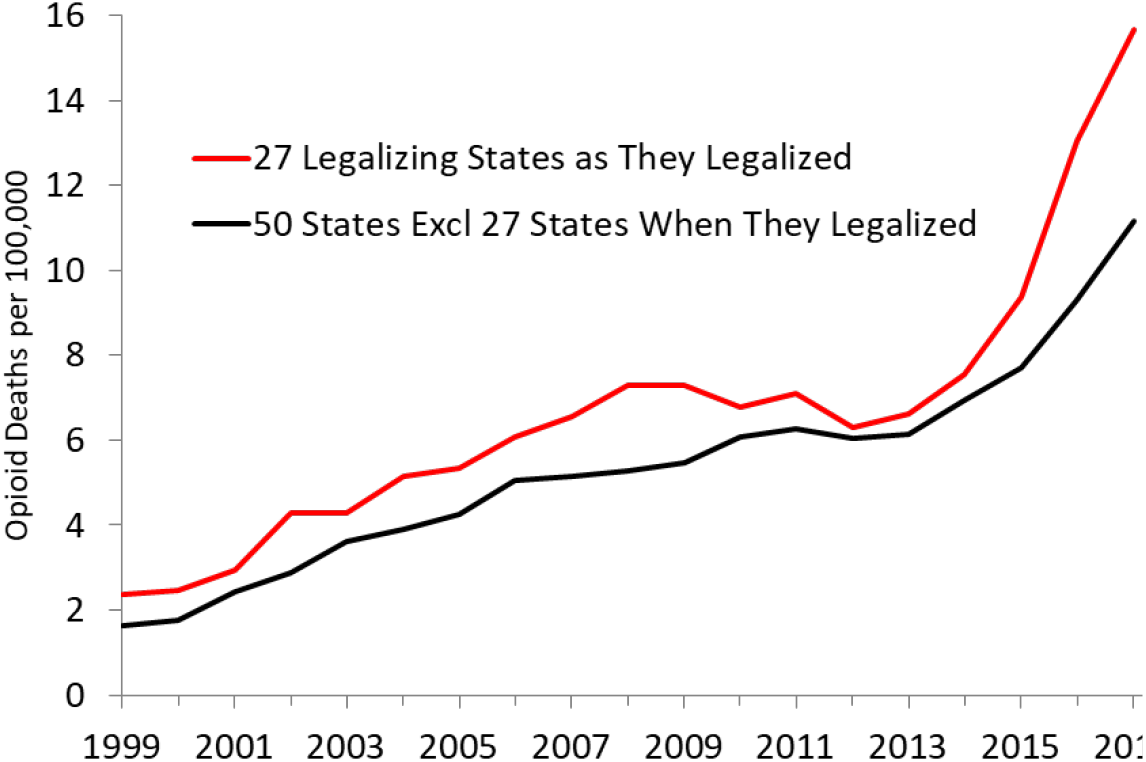
Annual Opioid Death Rate of U.S. States, 1999-2017, by Marijuana Legalization Status. <u>Data Source</u>: Shover CL, Davis CS, Gordon SC, Humphreys K.^16^ Years with <10 deaths not included

**Figure 11.**
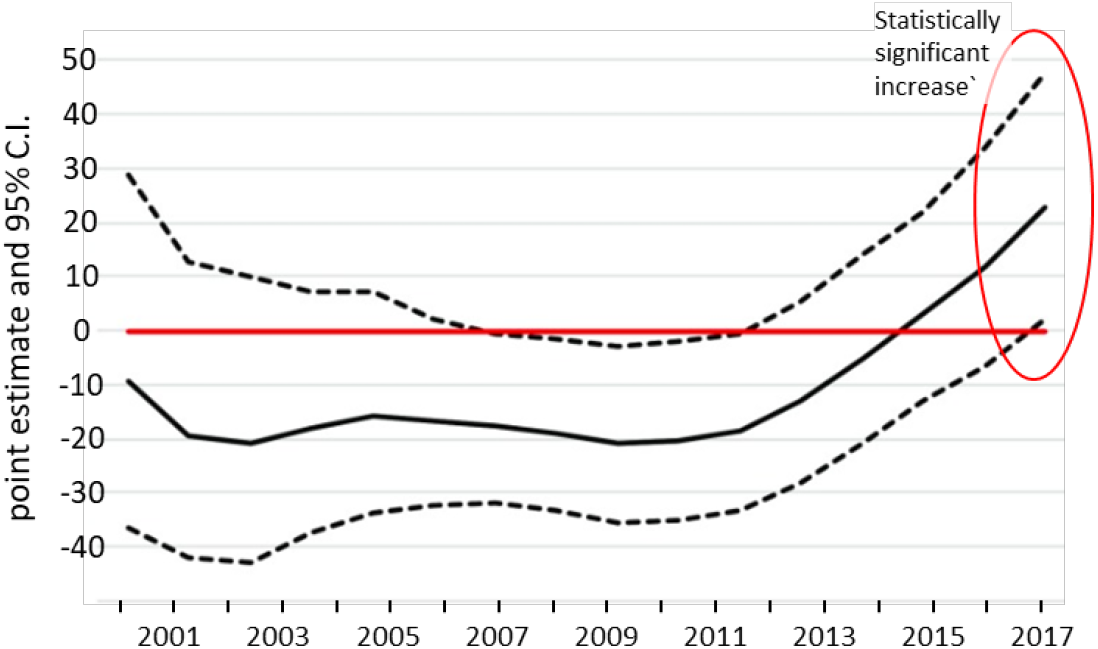
Annual Changes in Point Estimate and 95% CI of Association Between Medical Cannabis Law and Age-Adjusted Opioid Overdose Death Rate during 1999-2017. Fixed (year and state) and time-varying effects with adjustment for prescription drug monitoring program, state unemployment, pain management clinic oversight laws, and prescription drug identification laws. <u>Source</u>: Modified from Shover CL, Davis CS, Gordon SC, Humphreys K.^16^

Current evidence from the entire U.S., with more states, D.C. and longer follow-up than previously reported, does not support the “marijuana protection” hypothesis. If anything, the evidence presented here suggests the opposite, with 78% of the evaluable states and D.C. that legalized marijuana having opioid death rate trends consistent in greater increases after legalization than their pre-legalization rate or concomitant rate in the non-legalizing states. Collectively, since 2009-2011 when their rates were the same, the legalizing jurisdictions have had a highly statistically-significant greater increase in their opioid death rate compared to non-legalizing states. The results implicate the jurisdictions that implemented marijuana legalization as accounting for nearly three-fourths of the national opioid death rate during 2009 2017. Overall, the annual national opioid mortality rate was highly correlated with the cumulative proportion of the U.S. population in legalizing jurisdictions (p <10^−10^, Fig. 12).

**Figure 12.**
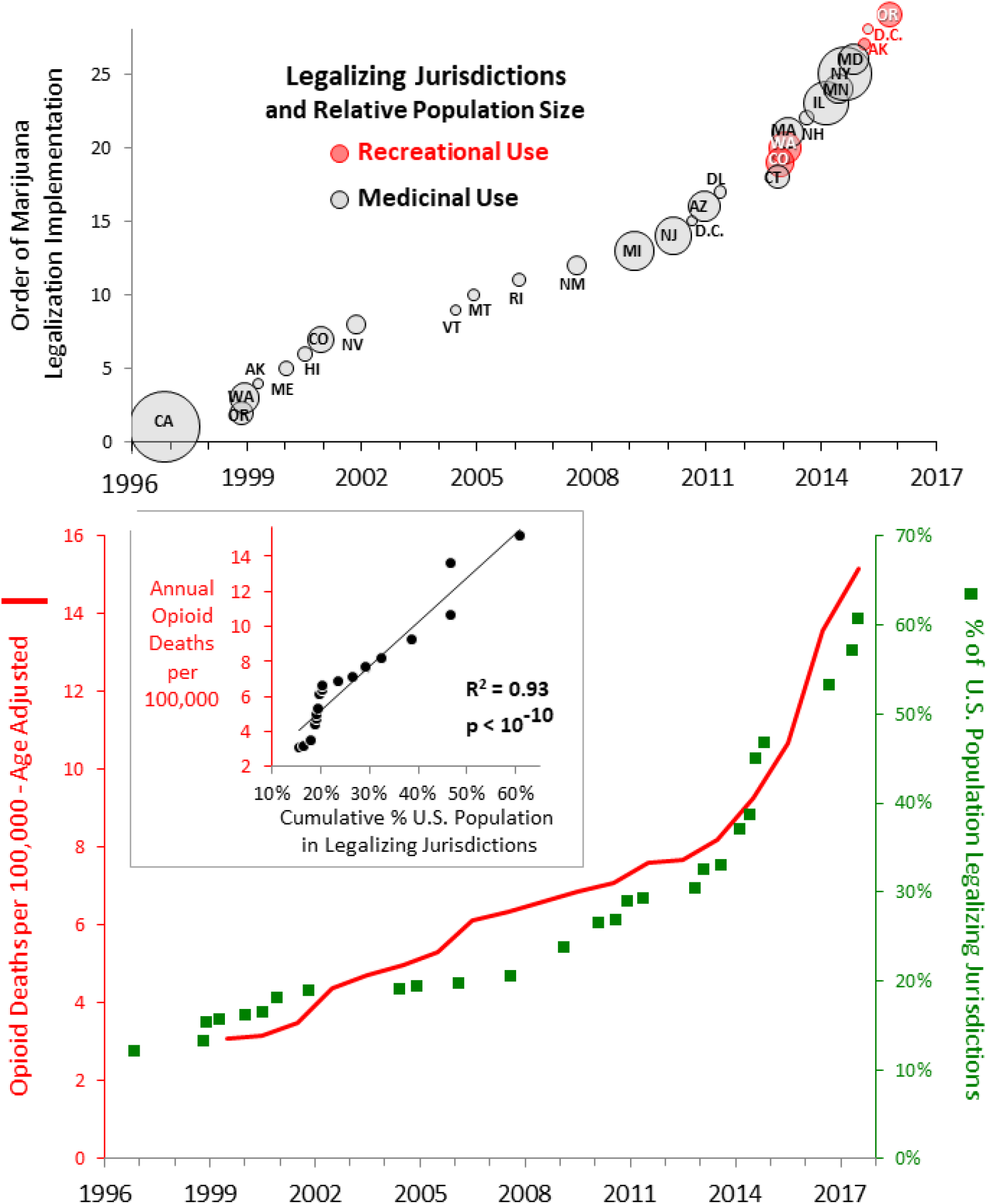
Order of Jurisdiction Legalization (upper panel) and Annual Cumulative Percent of U.S. Population in Marijuana Legalizing Jurisdictions and Opioid Death Rate (lower panel) in the U.S., 1996-2017, and Correlation of Opioid Death and Cumulative % of Population in Legalized Jurisdictions (lower panel inset). <u>Data Source</u>: CDC WONDER^2^

Our investigation has several limitations. Most important, the ecologic design does not establish attribution of causation. Other potential contributing factors other than marijuana legalization that may have resulted in the marijuana legalizing jurisdictions having a higher opioid death rate include strengthening of prescribing laws and regulations in non-legalizing states and the reduction of opioid supply in 2010-2012 and the invasion of illicitly manufactured fentanyl and fentanyl analogs that started in 2014-2015 that may have selectively accelerated the transition to heroin in the legalizing jurisdictions. As also noted by Bachhhuber et al,^3^ we could not adjust for differences between states in socioeconomic status, race/ethnicity, or medical and psychiatric diagnoses that may have caused more opioid deaths in the legalizing jurisdictions.

We also could not assess state level variation in the availability of semisynthetic narcotics (fentanyl and analogues)^17^ and of gabapentinoids increasingly being used to potentiate the euphoric effect of opioids.^18^ Nor did we assess federal vs. local governmental drug policy, funding, and treatment availability. As emphasized by Pacula et al,^19^ we did not account for state-level access to marijuana due to differences in medical laws across states (qualifying conditions, presence and legal protection of dispensaries, etc.) and recreational laws (passage vs. implementation dates, retailer density, price). With only four evaluable recreational-legalizing jurisdictions, potential differences in the impact of medicinal and recreational legalization could not be quantitated, albeit in the U.S. the degree of overlap between medicinal and recreational cannabis users has been estimated to be 86%.^17^ Death certificate data may have not correctly classified cases of opioid deaths. With only four evaluable recreational-legalizing jurisdictions, potential differences in the impact of medicinal and recreational legalization could not be quantitated, albeit in the U.S. the degree of overlap between medicinal and recreational cannabis users has been estimated to be nearly 90%.^19^ We did not use difference-in-difference, interrupted time-series design, or synthetic cohort methodologies applied in prior reports^3-5^ since the legalizing cumulative aggregate and non-legalizing group of jurisdictions had the same rates for three consecutive years during 2009-2011 and statistically similar rates the next year, 2012.

The current investigation also has several advantages over the prior reports. It assesses all states, includes D.C., and evaluates each legalizing jurisdiction both individually and collectively in a cumulative composite aggregate. The individual jurisdiction evaluations include comparisons of post-legalization and pre-legalization trends, comparison of post legalization and non-legalizing jurisdiction composite trends, and both of the aforementioned comparisons. It adds 16 states, D.C., and seven additional follow-up years to the Bachhuber study,^3^ 10 more states and four more years to the Powell study,^4^ and two more years to the Livingston srudy.^5^ It evaluates medicinal and recreational legalization effects, both separately and together, whereas both the Bachhuber and Powell studies only assessed medicinal legalization. It evaluates death rates, age-adjusted for annual rates, whereas Livingston et al^5^ assessed only deaths that were not age-standardized. Although we did not use the difference-in-difference methodologies, our selection of joinpoint analysis has the advantages of direct pairwise comparison and identifying when trend changes occur, the probability range of the inflection, and without requiring parallel trend assumptions. We included the T40.6 category of semi-synthetic opioids (including fentanyl and analogues), which had not been coded prior to October 2014^20^ and therefore not assessed by Bachhuber et al^3^ or Powell et al^4^. We included heroin (T40.1) and opium (T40.0) whereas Bachhuber et al^3^ did not. (Livingston et al^5^ did not specify codes they used).

In retrospect, the prior studies were premature assessments. The last year assessed by Bachhuber at al^3^ was 2010 and by Powell et al^4^ it was 2013. Livingston et al^5^ reported a “reversal of the upward trend in opioid-related deaths” after recreational legalization in Colorado, but assessed the post-legalization trend for just 24 months. Our analysis covers 4 years after recreational legalization during which there is no evidence of a decline in the state’s opioid death rate (Fig. 4 upper panel), nor in a 2019 report from the Oregon Department of Public Health and Environment.^21^

If our results indicate that increased availability of marijuana increases opioid use, abuse or overdose potential, this interpretation is consistent with other recent observations. A study conducted by investigators at the National Institute on Drug Abuse, New York State Psychiatric Institute and Columbia University Medical Center concluded that within 3 years of using cannabis, nonmedical prescription opioid use increased 5.8 fold (95% CI=4.2-7.9) and opioid use disorder increased 7.9 fold (95% CI=5.0-12.3).^22^ A 4-year prospective cohort study of 1,514 patients with chronic non-cancer pain, in whom 24% used cannabis for pain, those who used cannabis daily or near-daily used more opioids than those who did not (from their data we calculated a 39% higher oral morphine equivalent consumption, p=0.0001).^23^ In the an individual-level analysis of a nationally representative sample, medical cannabis use was positively associated with greater use and misuse of prescription opioids.^24,25^ In another study, self-reported marijuana use during injury recovery was associated with an increased amount and duration of opioid use.^26^ Conversely, medical prescriptions for Medicaid and Medicare Part D patients have been reported to be associated with reductions in opioid prescribing after legalization.^27-30^ A reduction of medical prescriptions for opioids may, however, increase the pursuit of black market opioids of uncertain composition and greater risk.^31^

Behaviorally and socially, marijuana may be a gateway to the use and eventual abuse of opioids and other addicting substances.^32-35^ Biologically, a gateway explanation for the marijuana-opioid connection is plausible since cannabinoids act in part via opioid receptors^36,37^ and increase dopamine concentrations in the nucleus accumbens similarly to that of heroin and related drugs with abuse potential.^36,38^ Clinically, cannabis use may reduce opioid withdrawal symptoms as a “low-efficacy” agent^39,40^ but to date, no prospective evidence, either from clinical trials or observational studies, has demonstrated any benefit of treating patients who have opioid addiction with cannabis.^25^ Cannabis’ ameliorating effect may also alleviate opioid withdrawal symptoms and thereby allow opioid use. Legalizing jurisdictions may have a culturally greater affinity for substance use disorder in general and be more vulnerable to gateway mechanisms. Although legalization is expected to decrease illicit activity, the black market may paradoxically benefit from access to more abundant crops, provide lower prices since none of the legalizing jurisdictions have regulated cannabis products,^41^ and by delivering cannabis to users instead of them having to travel to licensed dispensaries.

Trends of individual legalizing jurisdictions and their composite suggest that, if causally related, it initially took several years for legalization implementation to accelerate the opioid mortality rate. As legalization accelerated in the U.S., however, the interval to opioid mortality acceleration appears to have shortened to two years or less (Fig. 12).

A variety of suggestions to reverse the opioid death epidemic have been offered.^31,42-66^ In 2017, the National Academy of Sciences concluded that the myriad of studies of the public impact of cannabis use in all its various forms have not appropriately synthesized, translated for, or communicated to policy makers, health care providers, state health officials, or others responsible for influencing and enacting policies, procedures, and laws related to cannabis use.^67^ A more recent review concluded that recent state regulations that allow medical cannabis as a substitute for opioids for chronic pain and for addiction has at best equivocal evidence regarding safety, efficacy, and comparative effectiveness.66 The investigators concluded that substituting opioid addiction treatments with cannabis is potentially harmful and does not meet standards of rigor desirable for medical decisions.^68^

Meanwhile, marijuana use and legalization continues to expand. This direction in the U.S., when it leads the world both in cannabis use disorder and in opioid mortality, merits considering marijuana legalization as a contributing factor.

## Conclusions

Population data in the country with the highest prevalence of cannabis use disorder do not support the marijuana protection hypothesis and may indicate the opposite. Recommendations to enact laws to allow medical or recreational cannabis use should not be based on attenuating the opioid crisis. Before other states and countries “rush” to legalize marijuana and risk worsening the opioid crisis, the marijuana-opioid interaction should be more definitively researched.

## Data Availability

The global data are from the Institute for Health Metrics and Evaluation (IHME). GBDCompareDataVisualization. Seattle, WA: IHME, University of Washington, 2016. Available from http://vizhub.healthdata.org/gbd-compare. (Accessed October 22, 2018).
U.S. data not compared with other countries are from the Centers for Disease Control and Prevention, National Center for Health Statistics. Multiple Cause of Death 1999-2016 on CDC WONDER Online Database, released December 2017. Data are from the Multiple Cause of Death Files, 1999-2016, as compiled from data provided by the 57 vital statistics jurisdictions through the Vital Statistics Cooperative Program.

https://wonder.cdc.gov/ucd-icd10.html

## Acknowledgements

The authors wish to acknowledge the stress and harm to families and communities caused by addiction. We expect that science-driven clinical understanding and governmental actions will ultimately reduce this burden.

